# Effect of COVID-19 vaccination on menstrual periods in a retrospectively recruited cohort

**DOI:** 10.1101/2021.11.15.21266317

**Authors:** Victoria Male

## Abstract

Surveillance schemes are receiving increasing numbers of reports from people who have noticed a change to their period following COVID-19 vaccination. In order to investigate this, we retrospectively recruited 1273 people who have a record of their menstrual cycle and vaccination dates and used their reports to explore hypotheses about how COVID-19 vaccination and menstrual changes could be linked. In this dataset, we were unable to detect strong signals to support the idea that COVID-19 vaccination is linked to menstrual changes. However, larger, prospectively recruited studies may be able to find associations that we were not powered to detect.

## Introduction

As the UK COVID-19 vaccination programme is rolled out to younger participants, the MHRA’s surveillance scheme, Yellow Card, is increasingly receiving reports from people who have noticed a change in their menstrual cycle following vaccination: at 10 November 2021, 37571 such reports had been made [1]. It is important to note that most people who report such a change following vaccination find that their period returns to normal the following cycle [2] and that there is no evidence that COVID-19 vaccination adversely affects female fertility [3 - 7]. Nonetheless, people are concerned by these reports. Investigating the potential link between COVID-19 vaccination and menstrual changes is important for maintaining public trust in the vaccination programme and, if a link is found, to allow people to plan for potential changes to their cycles [8].

We currently know very little about how vaccination may affect the menstrual cycle, but there is some evidence that HPV vaccination may be associated with heavier or irregular periods [9]. There is also evidence that viral infection, including with SARS-CoV2 itself, can alter the menstrual cycle [10, 11]. This may suggest that immune stimulation can affect the menstrual cycle. Biologically plausible mechanisms by which this could occur include effects mediated by immunological influences on the hormones driving the menstrual cycle [12] or by immune cells in the lining of the uterus, which are involved in the cyclical build-up and breakdown of this tissue [13].

To address the question of whether there is a link between COVID-19 vaccination and changes to the menstrual cycle, we have recruited two cohorts. The first is a prospectively recruited cohort of 250 people, who are tracking their menstrual cycles before and after COVID-19 vaccination. Data collection from this cohort is still ongoing, but we expect that this will give us some idea of how commonly menstrual changes occur following vaccination. Here, we provide a preliminary analysis of a second cohort, recruited retrospectively, of 1273 people who keep a record of their menstrual cycles and vaccination dates. Because the cohort is likely to be enriched for people who noticed a change to their cycle, we cannot use this data to estimate how common postvaccination menstrual changes are. However, we can use the data to test some hypotheses about how COVID-19 vaccination and menstrual changes may be connected. This may give us a first idea of whether there is, indeed, a causal link between COVID-19 vaccination and menstrual changes.

## Methods

We recruited 2241 people who were over 18, had received at least one dose of a COVID-19 vaccination, have periods or withdrawal bleeds and who have a record of the dates of their periods, and the date or dates on which they received the vaccine. We asked them to use a web-based form to anonymously report their age, length of their normal menstrual cycle, whether they use any hormonal contraception, whether they are breastfeeding, whether they have ever been diagnosed with a menstrual or gynaecological condition and, for each dose of the vaccine, which brand they had, on which day of their cycle they were vaccinated and details of how the timing and flow of their next period compared to what they normally experience. Ethical approval for this data collection was given by the Research Governance and Integrity Team at Imperial College London, study number 21IC6988.

Before examining the data, we pre-specified the analysis plan https://osf.io/pa3nd.

The following responses were cleaned from the data set: 0. Responses made in preview mode (survey tests by the study team and for ethical approval; n = 7). 1. Responses in which the normal cycle length was not given (n = 33). 2. Responses where the normal cycle length was given as less than or equal to 19 days; the majority of these responses were in the range of 3-7 days, suggesting that respondents may have given the length of their menses, rather than their menstrual cycle (n = 646). 3. Responses where the normal cycle length was given as more than or equal to 40 days (n = 25). 4. Responses where the normal cycle length was given by a range, where the range was >3 days (n = 36). 5. Responses where there was a text indication that cycle length was unpredictable or irregular (n = 38). 6. Responses in which the day of the cycle that the vaccine was given was not specified (n = 140). 7.

Respondents who did not have a period after the vaccine, but the reason is specified (eg. became pregnant in the same cycle that they were vaccinated, opted not to have a withdrawal bleed if taking a contraceptive pill; n = 28). 8. People who did not receive the vaccine (n = 2). 9. One respondent noted in the freetext box that they had made an error, and would resubmit a fresh version (n = 1). 10. Respondents who noted that they do not normally have periods or withdrawal bleeds, not already removed from the dataset (n = 9).

After data cleaning, 1273 records remained, of which 813 had data for both the first and second dose of the vaccine. Demographic details of the respondents are given in Table 1.

**Table 1.** Characteristics of respondents.

| Demographic characteristics |  |
| --- | --- |
| Age in years (Median, IQR) | 33 (29 – 39) |
| Cycle length in days (Median, IQR) | 28 (27 – 30) |
| Hormonal contraception |  |
| No hormonal contraception | 1117 (87.6%) |
| Combined pill | 53 (4.2%) |
| Progesterone only pill | 17 (1.3%) |
| Contraceptive implant | 2 (0.2%) |
| Contraceptive patch | 3 (0.2%) |
| Intrauterine system | 47 (3.7%) |
| Vaginal ring | 2 (0.2%) |
| Other | 10 (0.8%) |
| Not specified | 22 (1.73%) |
| Menstrual and gynaecological diagnosis |  |
| Abnormal menstrual bleeding | 22 (1.7%) |
| Heavy menstrual bleeding | 150 (11.8%) |
| Endometriosis | 60 (4.7%) |
| Polycystic ovaries | 87 (6.8%) |
| Uterine fibroids | 21 (2.4%) |
| Breastfeeding |  |
| No | 1179 (92.5%) |
| Yes | 89 (7.1%) |
| Not specified | 5 (0.4%) |
| Previous pregnancies |  |
| 0 | 572 (45%) |
| 1 | 219 (17.2%) |
| 2 | 220 (17.3%) |
| 3 or more | 252 (19.8%) |
| Not specified | 10 (0.8%) |
| Vaccine |  |
| AstraZeneca | 346 (27.1%) |
| Janssen | 8 (0.6%) |
| Moderna | 136 (10.7%) |
| Pfizer | 778 (61%) |
| Not specified | 5 (0.4%) |

Where a range was given for normal cycle length, the median cycle length was used for analysis. For examination of the effect of the day of the cycle on which the vaccine was given, the day of ovulation was estimated by cycle length – 14, based on the observation that the luteal phase of the menstrual cycle (after ovulation) is normally constant at around 14 days [14]. The day on which the vaccine was given, relative to the predicted day of ovulation, was therefore calculated by cycle day of vaccination – (cycle length – 14).

For our four pre-specified analyses, we tested independence between the following pairs of variables using Chi squared tests: 1a. Brand of vaccination and timing of next period; 1b. Brand of vaccination and flow of next period; 2a. Use of hormonal contraceptives and timing of next period; 2b. Use of hormonal contraceptives and flow of next period; 3a. Timing of vaccination and timing of next period; 3b. Timing of vaccination and flow of next period; 4a. Timing of period following dose 1 and Timing of period following dose 2; 4b. Flow of period following dose 1 and flow of period following dose 2. For analyses 1-3, data from both first and second doses were included. For analysis 4, records in which only first dose data was available were excluded.

In response to public interest, we added an exploratory analysis (not pre-specified) to determine if people who have already received a diagnosis of menstrual or gynaecological conditions are more likely to experience menstrual changes following vaccination. Data from both first and second doses were included and independence was tested using a Chi squared test.

A total of 10 statistical tests were carried out. The Holm-Bonferroni sequential correction was used to correct for multiple hypothesis testing. Tests in which the adjusted p valule (p’) is less than or equal to 0.05 are indicated and categories in which the standardised residual is greater than the critical value for residuals (1.96) are indicated by *. These categories have more responses in them than would be expected if the variables under investigation were independent.

## Results

### Brand of vaccine is not associated with differences in timing or flow of next period

Menstrual changes have been reported after receiving all brands of vaccines [1], suggesting that no particular brand or strategy (mRNA vs adenovirus-vectored) is clearly associated with menstrual changes. To confirm this finding within our dataset, we looked for associations between the proportion of respondents reporting a change in the timing (Figure 1a) or flow (Figure 1b) of the period following their vaccination, stratified by brand of vaccine received. The small number of respondents (n = 13) who had either received Janssen or did not specify were excluded from this analysis. In line with the reports made to the Yellow Card surveillance scheme, in this dataset there was no association between brand of vaccine received and self-reported change to timing or flow of the next period.

**Figure 1.**
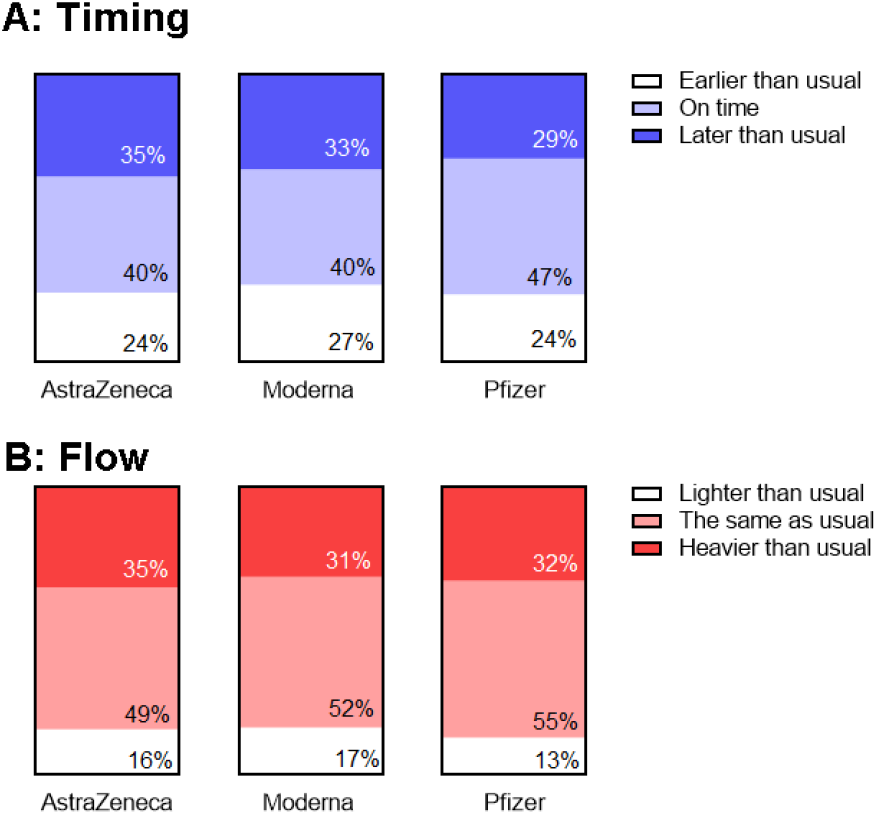
Examination of the association between the proportion of respondents reporting a change in the timing (A) or flow (B) of the period following their vaccination, stratified by brand of vaccine received.

### People on hormonal contraception were more likely to report a change to menstrual flow

If there is a link between COVID-19 vaccination and changes to periods, and it is mediated by changes to sex hormones, as has been suggested [12], then we might expect that people in whom exogenous sex hormones are supplied by hormonal contraception would be less likely to experience a menstrual change following vaccination.

To test this hypothesis, we looked for associations between the proportion of respondents reporting a change in the timing (Figure 2a) or flow (Figure 2b) of the period following their vaccination, stratified by whether the respondent was on hormonal contraception. Respondents who did not specify which form of contraception they use were excluded from the analysis. We found no association between hormonal contraception and timing of the next period but, contrary to our hypothesis, people on hormonal contraception were more likely to report that the flow of the period following vaccination was different from usual.

**Figure 2.**
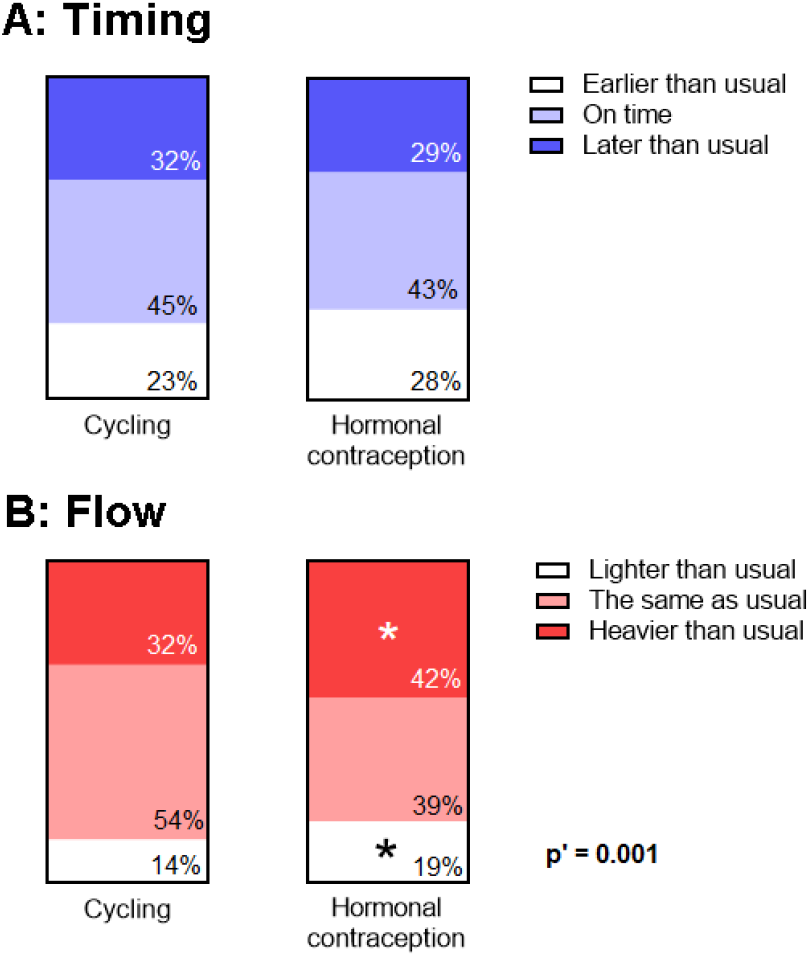
Examination of the association between the proportion of respondents reporting a change in the timing (A) or flow (B) of the period following their vaccination with use of hormonal contraception.

### Timing of vaccination does not have a clear effect on timing or flow of next period

One reason it has been difficult to use surveillance data to determine whether there is a link between COVID-19 vaccination and menstrual changes is that the reported changes have been very varied. However, we can postulate biologically plausible mechanisms by which varied changes might be linked to vaccination. For example, vaccination before ovulation might delay ovulation or prevent ovulation, lengthening the cycle. In this case, we might expect to see later than usual periods associated primarily with vaccination before ovulation.

To examine this, we stratified reports of the timing (Figure 3a) and flow (Figure 3b) of the next period depending on the day of the menstrual cycle on which the vaccine was given, relative to predicted day of ovulation. For this analysis, we examined only those who were not on hormonal contraception. We excluded menstrual cycle days on which fewer than 5 respondents had been vaccinated (excluding the responses for the 16 people vaccinated more than 17 days before the predicted day of ovulation) and days on which the respondent was already overdue for their period (n = 70), since these respondents would, by definition, report that their period was later than usual.

**Figure 3.**
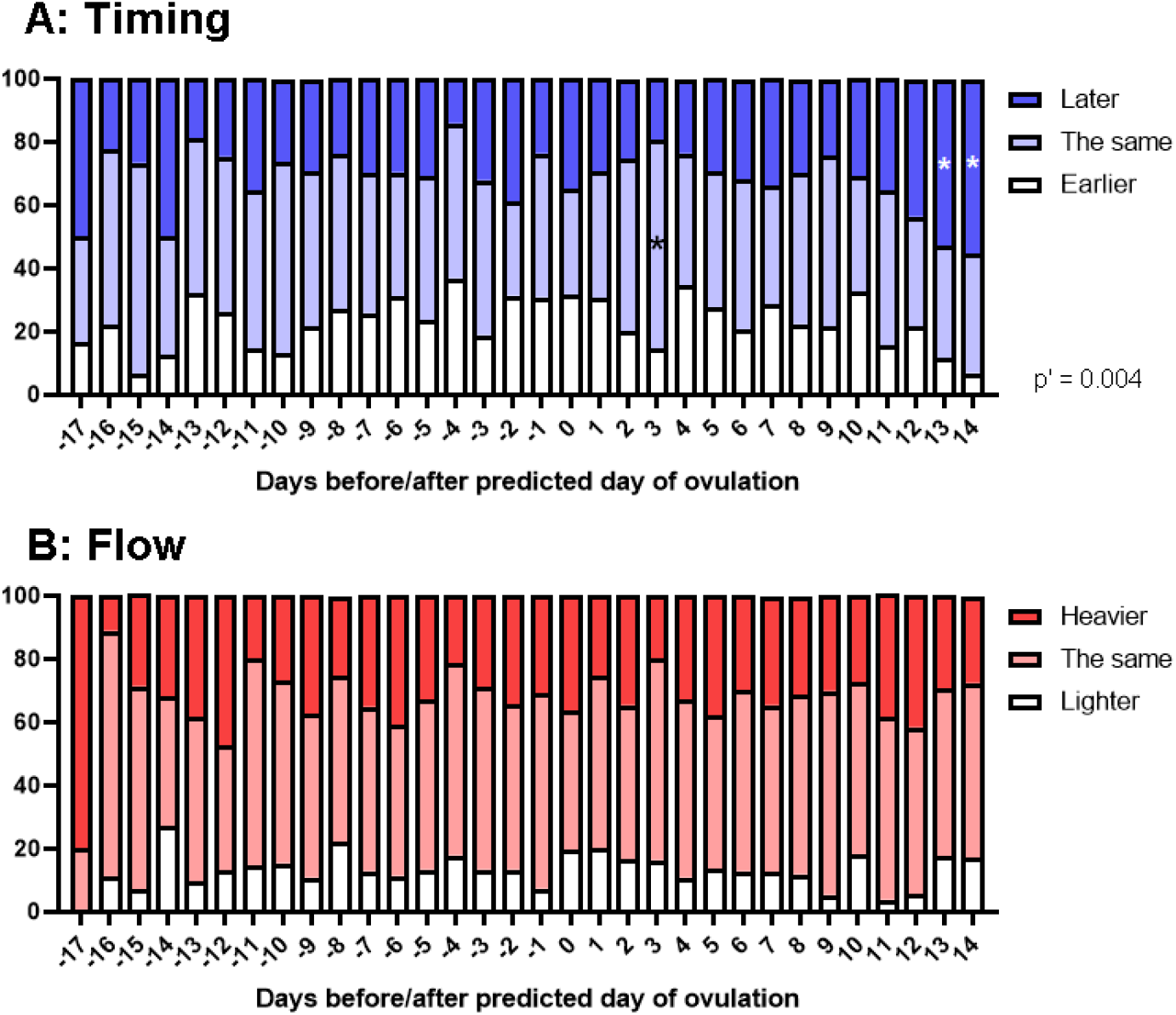
Reported timing (A) and flow (B) of the period following vaccination, stratified by day of the menstrual cycle on which the vaccine dose was given, relative to predicted day of ovulation.

We found a significant association between timing of vaccination within the menstrual cycle and the timing of the next period. However, examination of the standardised residuals revealed that this association was due to respondents vaccinated the day that their period was due, or the day before their period was due, who were more likely to report that their next period was late. This is perhaps unsurprising, since the timing of vaccination meant that these people were already moving towards having a later than usual period. We found no association between timing of vaccination and flow of the next period.

### Menstrual changes following dose 2 are closely associated with those following dose 1

Another possible explanation for the variety of reported menstrual changes following COVID-19 vaccination is that individual or genetic factors may affect the kinds of changes that people experience. If this were the case, we would expect changes experienced following dose 1 to predict those following dose 2.

To examine this possibility, we looked at whether reported timing (Figure 4a) and flow (Figure 4b) of the period following dose 2 was associated with the report following dose 1. For both timing and flow, we found a strong association between the reports for dose 1 and dose 2, with people most likely to report the same experience following dose 2 as they had following dose 1. This could support the idea that there is some interindividual variation in menstrual changes following COVID-19 vaccination, but it is also important to consider the other possible reasons for this relationship (see Discussion).

**Figure 4.**
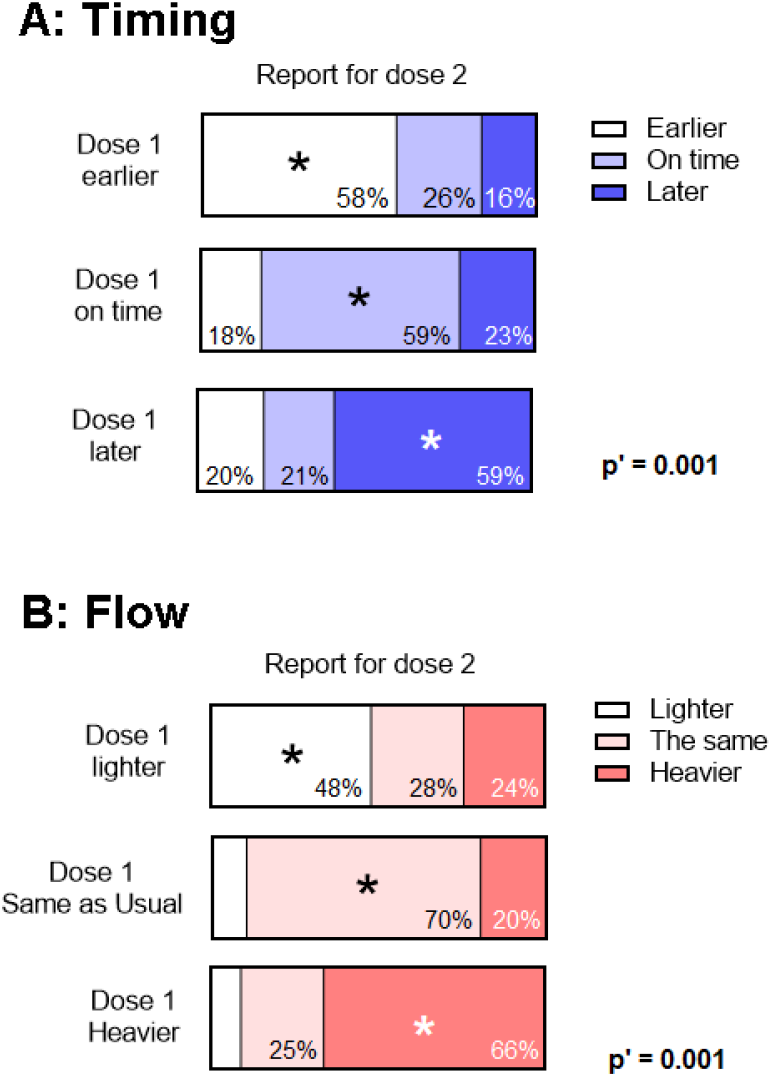
Association between reports of changes in timing (A) and flow (B) following first and second vaccine dose.

### People who have a diagnosis of endometriosis or PCOS may be more likely to experience a change in the timing of their cycle following vaccination

A number of people have approached us with the concern that, since they already experience heavy or otherwise difficult periods because of their pre-existing conditions, any menstrual changes following COVID-19 vaccination might be more pronounced for them. Our conversations suggest that this is a major contributor to vaccine hesitancy in this group.

To address this concern, we added an exploratory analysis (not pre-specified) to look at how having a pre-existing diagnosis of a menstrual or other gynaecological condition might affect the timing (Figure 5a) or flow (Figure 5b) of the period following vaccination. We did not observe any association between reported flow and having had a diagnosis of abnormal menstrual bleeding, heavy menstrual bleeding, endometriosis, polycystic ovaries or uterine fibroids, or no reported diagnosis. After correction for multiple hypothesis testing, the association between the timing of the next period and pre-existing diagnosis was borderline significant (p’ = 0.05). People with a diagnosis of abnormal or heavy menstrual bleeding or uterine fibroids did not differ in their reports from those with no diagnosis, but those with a diagnosis of endometriosis were somewhat more likely to report an earlier than usual period, and those with a diagnosis of polycystic ovaries were somewhat more likely to report a later than usual period.

**Figure 5.**
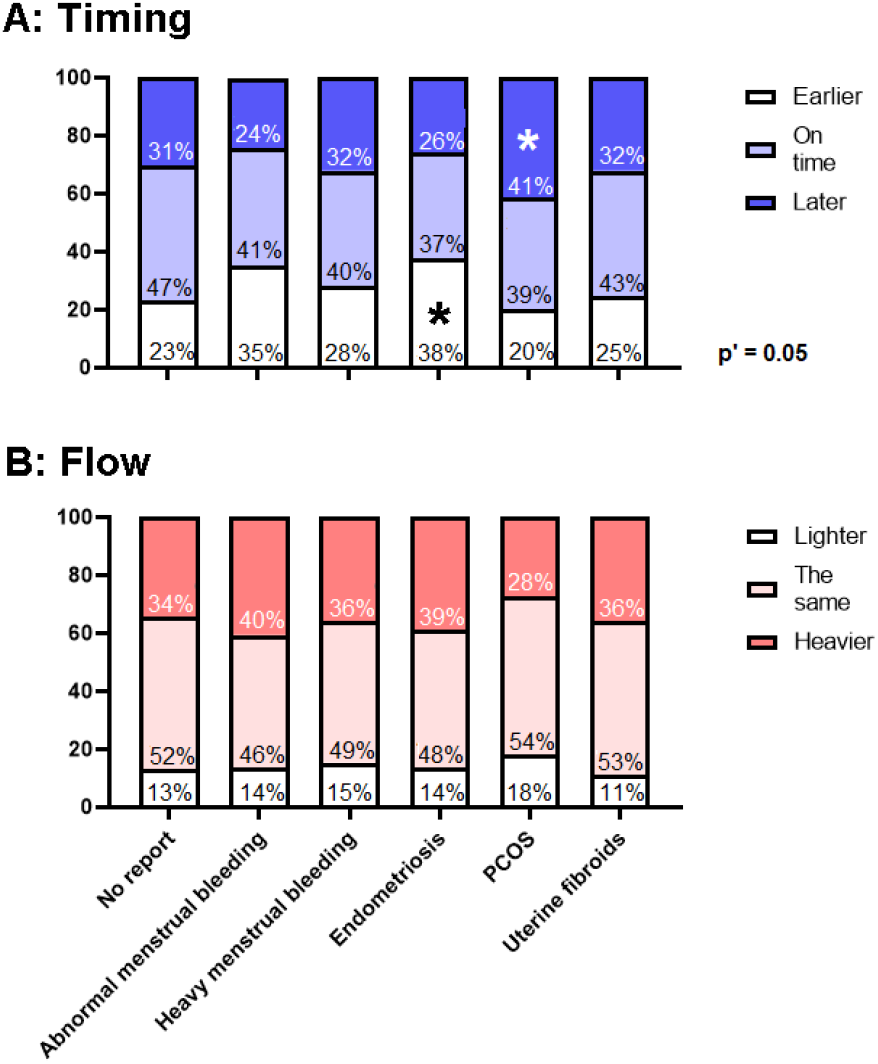
Examination of the association between the proportion of respondents reporting a change in the timing (A) or flow (B) of the period following their vaccination with pre-existing diagnosis of a menstrual or gynaecological condition.

## Discussion

Here, we report on the experiences of 1273 people, recruited retrospectively, who kept a record of their periods and the date of their COVID-19 vaccination. Our findings suggest that there is no association between brand of vaccine and changes to periods. The association we identified between having a dose of COVID-19 vaccine in the last two days of the menstrual cycle and the subsequent period being later than usual is likely to be accounted for by the period or withdrawal bleed in these cases already being late at the time of vaccination.

Unexpectedly, we found that people using hormonal contraception were more likely to report a change to the flow of the period immediately following COVID-19 vaccination. This finding opposes the idea that any change to vaginal bleeding following COVID-19 vaccination is mediated by changes to hormones, since in this case we would expect those on hormonal contraception to be less likely to experience changes. It is difficult to propose a biological reason that those on hormonal contraception should experience more menstrual changes than those who are naturally cycling, so we should consider the possibility that this finding may be a result of reporting bias. Many people use hormonal contraception at least partially to make their bleeds lighter and more regular [15,16], so people on hormonal contraception who experienced a change may have been more motivated to respond to the survey.

We found that people’s reports of the timing and flow of their period following first vaccination dose was strongly predictive of their report following second vaccination dose. This could be consistent with interindividual factors affecting the nature of the menstrual response to vaccination, but there are other potential explanations for this observation. In this largely UK-based cohort (n = 1121, 88%), vaccine doses would have been given at around 8 weeks apart [17], meaning that any change, whether vaccine-related or otherwise, that affected the post-dose 1 period could potentially still be in effect for the post-dose 2 period.

We found that respondents who had a diagnosis of a menstrual or gynaecological condition were not more likely to report a change in flow than those who did not have such a diagnosis, and that those with a diagnosis of heavy or abnormal menstrual bleeding or uterine fibroids were not more likely to report a change in timing. We hope that our findings will be reassuring to people with these conditions. However, we did find a slight increase in the frequency of people with endometriosis who reported an earlier than usual period, and in people with polycystic ovaries who reported a later than usual period. It will be important to follow up this finding to determine whether these groups really are more likely to experience a change to the timing of their cycle. In the interim, we emphasise that these findings should not be used to counsel people who have these diagnoses against vaccination. Indeed, it is important for those who are particularly concerned about changes to their menstrual cycles to be reminded that COVID infection itself may cause this [10, 11].

The study has a number of limitations. First, because the participants were recruited retrospectively, the data is likely to be enriched for those who noticed a change, who might be more motivated to participate in the study. Therefore, we cannot use this data to determine the frequency with which people experience menstrual changes following COVID-19 vaccination or, directly, to confirm or disprove any link between vaccination and menstrual changes. Approaches in which participants are recruited prospectively or using menstrual cycle data collected for other reasons, for example, datasets from menstrual cycle tracking apps with linked data about dates of vaccination, are better equipped to answer these questions. Approaches using menstrual cycle tracking apps are likely to be particularly powerful because the large number of cycles logged and the granularity of the data will allow detection of small and rare changes to post-vaccination menstrual cycles. Further, where the app uses user data to predict the day of ovulation, the date of vaccination relative to ovulation can be determined with greater accuracy than the crude estimate used here.

It is also important to note that the majority of the participants in this study were from the UK, so our findings here might not be applicable to other countries. In particular, we did not examine any potential associations with vaccines that are not approved for use in the UK, such as Sinovac, Sinopharm or Sputnik V. In the UK, we use an interdose interval of at least 8 weeks, whereas most other countries use a 3- or 4-week interval, and this could also mean that different effects may be seen in countries vaccinating with these schedules, even when they are using the same vaccines.

In conclusion, this study of 1273 retrospectively recruited participants was unable to detect strong signals to support the idea that COVID-19 vaccination is linked to menstrual changes. However, large, prospectively recruited studies may be able to find associations that we were not powered to detect.

## Data availability

Cleaned, fully anonymised data, together with our analysis files, are available from the Open Science Framework at https://osf.io/6jf4u/.

## Supporting information

STROBE checklist for observational studies

## ACKNOWLEDGEMENTS

This study did not receive any funding. The investigator’s salary was paid by the Wellcome Trust (WT105677; to July 2021) and the preterm birth charity Borne (from August 2021).

